# A Qualitative Exploration of Patients’ Experiences of Living with Chronic Respiratory Diseases before and after Participating in a Pulmonary Rehabilitation Program at a Tertiary Hospital in Malawi, and Their Suggestions to Improve a Future Program

**DOI:** 10.1101/2023.12.14.23299953

**Authors:** Fanuel M. Bickton, Talumba Mankhokwe, Beatrice Chavula, Emily Chitedze, Martha Manda, Cashon Fombe, Martha Mitengo, Langsfield Mwahimba, Moses Isiagi, Richard N. van Zyl-Smit, Susan Hanekom, Martin Heine, Harriet Shannon, Jamie Rylance, Enock Chisati, Stephen B. Gordon, Felix Limbani

## Abstract

**Background:** Community-based surveys suggest a substantial burden of chronic respiratory diseases (CRDs) in the Malawian population, causing significant morbidity and loss of economic productivity. Pulmonary rehabilitation (PR) is an effective non-pharmacological intervention for people with CRDs, but there is limited data on its feasibility and acceptability in Malawi.

**Objectives:** To explore the experiences of patients with CRDs before and after participating in a PR program at Queen Elizabeth Central Hospital (QECH), Blantyre, Malawi, and their suggestions to improve a future program.

**Methods:** Fourteen adult patients (eight females and six males) presenting with functionally limiting CRDs to QECH were invited to participate in a six-week PR program. Following program completion, face-to-face semi-structured in-depth interviews with the participants were conducted. Interviews were audio-recorded and transcribed verbatim. The transcripts were anonymised and thematically analysed using a deductive approach.

**Results:** Ten adults (five females and five males) participated in the PR program. Their documented CRD diagnoses included chronic obstructive pulmonary disease (COPD), asthma, post-tuberculosis lung disease, and bronchiectasis. Five key themes emerged: (1) experiences of living with a CRD before PR, (2) experiences of living with a CRD after PR, (3) feedback on the conduct of the completed PR program, (4) suggestions to improve a future PR program, and (5) program continuation/maintenance at home. Participants reported experiencing improvements in physical, psychological, and social health associated with PR program participation. The provision of transport was considered a key facilitator for PR program completion. Realising the gained PR benefits, participants were willing to continue exercising at their homes, albeit with potential barriers including a lack of equipment.

**Conclusion:** The PR program improved the participants’ perceived health status and was well-accepted. Addressing barriers related to transport facilitated immediate implementation while providing a challenge for the scaling and sustainability of PR beyond the project duration. These findings support the drive for shifting chronic care, including rehabilitation, towards primary care and community.

**Trial Registration:** Prospective; 27/08/2021; ISRCTN13836793

## Introduction

For people with chronic obstructive pulmonary disease (COPD), pulmonary rehabilitation (PR) is supported by high-quality evidence of improvement in symptoms (dyspnoea, fatigue, anxiety, depression), exercise tolerance, and overall health-related quality of life (1). There is also evidence supporting PR for other chronic respiratory diseases (CRDs) including interstitial lung disease (2), pulmonary hypertension (3), asthma (4), cystic fibrosis (5), bronchiectasis (6), lung cancer (7), lung transplant (8), post-TB lung disease (9), and post-COVID-19 (10). Evidence from high-income countries (HICs) suggests that PR significantly reduces the direct costs of COPD by decreasing healthcare system usage, particularly unplanned hospital admissions (11).

A systematic review of PR in Southern Africa, however, found few trials, and generally low-quality evidence for efficacy, with no published data from Malawi (12). Therefore, we conducted a trial of a PR program among patients with functionally limiting CRDs at Queen Elizabeth Central Hospital (QECH), Blantyre, Malawi. This paper reports the results of the post-PR qualitative study which aimed to explore patients’ experiences of living with CRDs before and after participating in PR the program, and their suggestions to improve a future program.

## Methods

Our published protocol, available elsewhere (13), includes detailed information about study design, participants’ sample size, recruitment process, eligibility criteria, and data collection. A summary is presented here.

### Trial setting

Healthcare facilities in Malawi are organised into primary, secondary, and tertiary levels, linked through a referral system. The primary level includes community health centres, the secondary level includes district hospitals, and the tertiary level includes four central hospitals two of which are in the Southern Region including QECH in Blantyre. QECH is Malawi’s largest tertiary hospital catering for all primary- and secondary-level facilities for the Southern Region. The hospital includes a Chest Clinic which serves outpatients with chronic cardiac and respiratory conditions (such as chronic heart failure and asthma, respectively) every Wednesday.

### Trial participants

A total of fourteen protocol-eligible (13) patients (eight females and six males, aged ≥18 years) presenting with functionally limiting CRD symptoms to the Chest Clinic at QECH were consecutively identified, invited, and consented (by TM, BC, MMi, MMa, and BC) to participate in a PR program at the QECH’s Physiotherapy Department. However, four did not participate: one male patient died of a cardiovascular complication, one female patient relocated to another city, and two patients (one male and one female) became unreachable. The remaining ten patients (six females and four males) participated in the PR program to completion and were all interviewed post-PR. Their socio-demographic and clinical characteristics are shown in table 1.

**Table 1:** Participants’ socio-demographic and clinical characteristics.

| Participant's number | Sex | Age ranges (in years) | Documented respiratory diagnoses in the participants' hand-held health records | Other documented diagnoses/comorbidities |
| --- | --- | --- | --- | --- |
| 1 | Female | In her 40s | Asthma | Hypertension |
| 2 | Female | In her 20s | Post-tuberculosis (post-TB) bronchiectasis | Human Immunodeficiency Virus (HIV) |
| 3 | Male | In his 40s | Asthma | None |
| 4 | Female | In her 60s | Asthma | None |
| 5 | Male | In his 50s | Chronic obstructive pulmonary disease (COPD), bronchiectasis | HIV |
| 6 | Female | In her 40s | Post-TB | HIV |
| 7 | Male | In his 80s | Post-TB, COPD | Cor-pulmonale, hypertension |
| 8 | Female | In her 40s | Post-TB, Asthma | HIV, depression |
| 9 | Male | In his 20s | Post-TB lung fibrosis | HIV, depression |
| 10 | Female | In her 50s | Asthma, community-acquired pneumonia (CAP) | HIV |

No participant had previously participated in PR. Each participant was given transport money to attend each PR session, covering their travel costs for both to and from the PR centre. The amount of the transport money differed from participant to participant due to their different locations or distance to travel and was equivalent to the total amount of transport money each participant would have to pay for public transport (e.g., minibus and/or motorcycle) to and from the PR centre. Each participant was also given an additional one thousand Malawian Kwacha as remuneration for their time. Refreshments such as drinking water were provided to participants at the PR sessions.

### Trial intervention

The PR program trial was conducted between February and May 2023. To make it locally appropriate, the PR program’s design incorporated the perspectives of participants of a formative qualitative study (14), some of whom also participated in the actual program. The PR program ran for six weeks, twice weekly, at QECH’s Physiotherapy Department. A team of six physiotherapists (TM, BC, MMi, CF, MMa, LM) and a nurse (EC) delivered the PR sessions, coordinated by the principal investigator (FMB). Due to limited physiotherapy gymnasium space and busier physiotherapists in the morning, all PR sessions were scheduled in the afternoon (from around 13:00). They were delivered in a group format and consisted of an hour of individualized circuit exercise training, with its intensity progression according to individual participant’s improvement over time. The circuit included exercises for both upper and lower limbs designed to improve aerobic endurance and muscle strength such as dumbbell lifting, stair climbing, treadmill walking, stationery cycling, and sit-to-stand. Participants were encouraged to also exercise at their homes.

### Data collection

For an overview of quantitative measures included pre- and post-PR, we refer the reader to our published protocol (13).

Post-PR semi-structured in-depth interviews with individual participants were conducted by TM and MMi (both had prior qualitative research experience) using a topic guide (Table 2) which was informed by a literature review and received expert input from a Malawian social scientist (FL). It was not pilot tested but no significant areas for its quality or content improvement were identified as the interviews progressed. The interviews were conducted in the Head of Physiotherapy Department’s office, which was a quiet and private room within the Physiotherapy Department building at QECH, and their durations ranged from 10 to 38 minutes (22 minutes on average). All interviews were conducted in Chichewa (most spoken native language) and audio recorded.

**Table 2:**
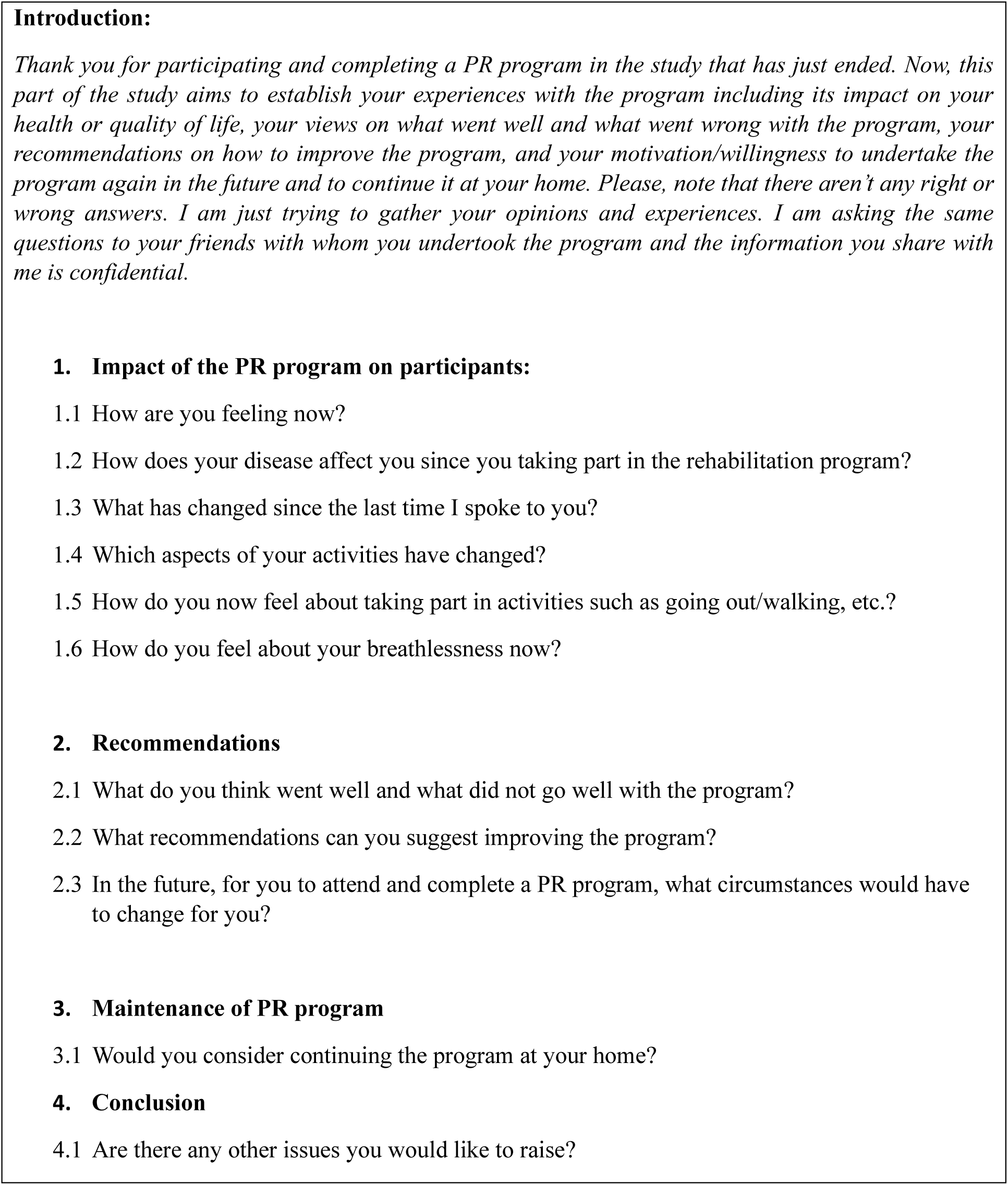
Semi-structured topic guide for the post-PR in-depth interviews.

### Data management

The interview audio files were stored on a password-encrypted computer, and transcribed verbatim (by FMB) in their original language (Chichewa) to minimize loss of meaning with English translation (15); only participant’s quotes included in this paper were translated in English at the time of results reporting. The interview data were anonymized at the time of translation and transcription by replacing participants’ names with a number (e.g., “participant 1”, “participant 2”, etc.).

### Data analysis

FMB and TM analyzed the qualitative data independently but collaboratively using a deductive thematic approach (16). Initially, the researchers collaborated to develop a preliminary coding framework. Data coding was performed manually by the researchers separately, using an iterative approach across all qualitative data (16). The researchers followed the steps outlined by Braun and Clarke (16), including independently going through a process of familiarization with the data by reading and re-reading transcripts while making reflective notes on the literal content, looking closely at words used by participants, interpreting what the data meant by assigning initial codes or classifications to segments of text, and exploring relationships between these classifications and developing core themes. The two researchers discussed their separate themes and merged them collaboratively.

### Ethics

Ethical approval for this study was obtained from the College of Medicine Research and Ethics Committee (COMREC), protocol number P.07/19/2752. Participants signed consent forms after willingly agreeing to participate in the study based on study information provided by the investigators.

## Results

From the analysis, five key themes emerged: (1) experiences of living with a CRD before PR, (2) experiences of living with a CRD after PR, (3) feedback on the conduct of the completed PR program, (4) suggestions to improve a future PR program, and (5) program continuation/maintenance at home. We now describe these five themes and include their sub-themes and select illustrative participants’ quotes in tables 3–7, e.g., P1F40s is participant 1, female, in her 40s, etc.

### Theme 1: Experiences of living with a CRD before PR

Participants described various symptoms of their CRDs, which they said were more severe before they participated in the PR program. These symptoms included breathlessness, (productive) cough, wheeze, chest tightness, chest pain, dizziness, leg pain, a feeling of body heaviness, and body weakness. Before PR, these symptoms affected participants’ physical, psychological, and social health negatively as well as disturbed their sleep. The symptoms were a reason for some participants’ frequent use of inhaler, hospital visits and hospitalisations, although they felt that these were less helpful.

**Table 3:** Illustrative quotes about living with a CRD before PR.

| **Sub-theme** | **Illustrative Quote** |
| --- | --- |
| Symptom occurrence | *“Upon waking up in the morning, the body used to feel heavy, feeling like sick from malaria and what... to walk, [I used to feel] chest pain, leg pain.” (P2F20s)* |
| Physical health | *“Before I started PR, when I walk – I like walking a lot – I used to experience frequent breathlessness. ... I used to stop frequently to rest.” (P5M50s)* |
| Psychological health | *“The difference [between pre- and post-PR] is that we used to be sedentary [before PR]. We were afraid of doing an activity that we could do, saying “If I do this with force, maybe it will worsen my condition.”” (P4F60s)* |
| Social health | *“Often, I could only be in one place, struggling with sickness, just sleeping. When my friends were going to parties or to church, I was failing to go because of sickness...” (P1F40s)* |
| Sleep health | *“... [before PR], my breathing used to be difficult... difficult especially at night when I sleep. When I sleep, and start coughing, I used to get very breathless, sometimes sweating. So, I could wake up [and] sit down, to do what? To rest, to get back my breath.” (P6F40s)* |
| Medication, hospital visits and hospitalisation | *“So, I used to come to the hospital frequently. Getting admitted in 4A [ward], sometimes [getting admitted] up there where there are lots of window glasses [AETC]; I used to be hospitalised there for a long time. Maybe wondering, “What is happening? I can’t see anything [the treatment received there] helpful.” Giving me Jackson [injection], giving me drugs, but nothing was what? Helpful.” (P8F40s)* |

### Theme 2: Experiences of living with a CRD after PR

Participants reported positive impact of the PR program on various aspects of their lives including improving their CRD symptoms. For example, they reported reductions in the magnitude and frequency of the symptoms, accompanied by improvements in their physical, psychological, social, and sleep health. In addition, participants reported acquisition of new and/or improved knowledge, including knowledge about exercise benefits. This knowledge helped them have a positive attitude towards exercises and/or physical activities, including removing their fear of engaging in them. Finally, due to PR, some participants reported using inhalers less frequently, the inhalers became more effective/responsive, and that they were visiting the hospital less frequently.

**Table 4:** Illustrative quotes about living with a CRD after PR.

| Sub-theme | Illustrative Quote |
| --- | --- |
| Symptom occurrence | *"What has changed... on the part of breathlessness, now I am not feeling as breathless as previously [before PR]. I am feeling [nowadays] breathless but occasionally, but I can do my household chores as a woman, being comfortable/relaxed. The breathlessness does not occur to me as frequently as previously." (P1F40s)* |
| Physical health | *"I am proud and happy because my body has now relaxed compared to how I was previously. Because even work, I was unable to do [before PR]; lifting [objects], I couldn't lift. But now, I can walk up a slight hill, on a flat surface too, walking well, or lifting and doing other household chores like laundry... doing household chores as a woman... while, previously, I couldn't do so." (P1F40s)* |
| Psychological health | *"... But when we I came here [after PR], I put aside many of the anxieties... they [anxieties] ended. ... I feel sweet in my life. It's good, it's done me good, coming here." (P8F40s)* |
| Social health | *"...these days [after PR], people are proposing me... But at that time [before PR], not even 'tsiii, tayimani' [Chichewa words to romantically stop a woman in the streets] ...was there. But these days, those with 'tsii' are? Increasing. But I think that all that power, or [my new] physical appearance, or the way they see me... because I did not have that power [to catch men's romantic attention] at that time [before PR], I didn't have enough strength, I couldn't walk smoothly or freely like one should do. But since I started this program [PR], I see that things for me have really what? Changed. Because I have the strength..." (P6F40s)* |
| Sleep health | *"Uhm, we can say that now it is better because, since the beginning of that program, any strange symptoms ... especially breathlessness, it's no longer happening or at night. Because sometimes... this breathlessness could come when I'm sleeping. Then, I would take the inhaler and puff it. But now, since this started [PR], I have not been breathless at night." (P5M50s)* |
| Knowledge | *Yes, but this [PR] program is really good because people were afraid that, "If I do that, I will be breathless. If I do this, I will be breathless", not knowing that maybe by doing so we are weakening our body a lot. But when we do this [PR], ... I have learned that activities are important ... a problem like that [breathlessness] can recover better if I do activities. Because many of us live in fear of danger, fearing that we might be breathless in this situation [of doing activities]. So, I have learned that it is important for the [PR] program to continue." (P10F50s)* |
| Medication, hospital visits and hospitalisation | *".... sometimes, I can do something without taking medicine; I am able to walk or climb, things that can still give us breathlessness, but finding that we are able to climb on that place... the breathlessness will come but it will not be necessary for us to use medicine. It's as if our bodies are relaxing, slowly relaxing because of those exercises when I take the medicine, I find that the medicine is responding quickly. Yes, that, "Ah, let's take her to the hospital", no, it [the exacerbation] resolves right there at home [without necessarily going to the hospital]." (P4F60s)* |

### Theme 3: Feedback on the conduct of the completed PR program

When asked about what went well and what did not, participants said the program went well generally. They attributed this to several reasons, including the afore-mentioned gained PR benefits, good personal attributes of the PR delivery team (such as welcoming, heartwarming/warmful, cheerful, kind, polite, hardworking, and participatory or engaging and interactive), transport money provision, and good exercise design (such as the group delivery format, exercise individualization, and the right choice of the exercises).

**Table 5:** Illustrative quotes of feedback on the conduct of the completed PR program.

| **Sub-theme** | **Illustrative Quote** |
| --- | --- |
| Personal attributes of the PR delivery team (such as welcoming, heartwarming/warmful, cheerful, kind, polite, hardworking, and participatory or engaging and interactive) | *“Truth is good. For me, from the first day [of PR] to come again today, everything was good. Yeah, because in this place, you didn't insult us ... but you were welcoming us as your children. Pleasant... anything was fun. Problems do not fail everywhere, but for me I saw that everything was a good thing.” (P7M80s)* |
| Transport provision | *“Yes, it [the PR program] also went well because you were providing transport money which made travel [to and from the PR centre] easier because according to the [financial] situation of people like us, maybe we should have started the program and left it on the way because sometimes we don't have money...” (P9M20s)* |
| Exercise design (group delivery, individualisation, exercise choice) | *“Every aspect has gone well. Yes, because... You were also looking at how sick a patient was, you were treating the patient according to how sick he/she was [exercise individualisation].” (P2F20s)* |

### Theme 4: Suggestions to improve a Future PR program

Participants suggested that, for more people with CRD to benefit from the PR program, the program should continue beyond the study, more patients should be recruited, and the program should be widely implemented in Malawi. Some participants suggested that this would be possible with further funding and government support for the program. In addition, participants suggested the need to raise awareness about the intervention, with themselves even willing to be champions of that awareness. Regarding timing of the PR sessions, some participants were okay with the afternoon sessions while some participants suggested shifting them from afternoon to morning hours for their convenience. Some participants’ suggestions were about adjusting the exercise design of the PR program which included increasing the intensity, diversity/variety, number, and duration of the exercise stations. They said these adjustments could help them benefit more from the program. Some participants suggested delivering comprehensive/structured education sessions within the PR program including education on safe home-based exercises and self-management skills.

**Table 6:** Illustrative quotes of suggestions to improve a future PR program.

| Sub-theme | Illustrative Quote |
| --- | --- |
| Program reach | *"... are you also going to reach other districts [with the PR program] or it's just around here, Blantyre, only?... if yes and people have been well-trained and realise its benefit, I hope that many can benefit. ... these [PR program] shouldn't just start and end, no, but should continue. Because, if you do it on a few people, there are many people who are suffering; maybe even we [current participants] are able to walk, we are coming here... others [with similar conditions] are indoors, they do not know how helpful physical exercises are. So, my opinions are that if the government agrees it [PR program] to be special that a person with such conditions as these should go and do physical exercises, not making us depend on drugs only, I feel like it might help. Of course." (P2F20s)* |
| Program awareness | *"I can say this: that those who can listen to me, listen ... maybe you can throw this [PR] program somewhere with the caption, "But so-and-so was helped. She is grateful for this [PR], she was helped. She was lacking good breathing, walking, and activity performance. But when she came to physiotherapy with this disease, it [PR] did her good." So that all can listen to a program like this, take a lesson from it, and be encouraged." (P8F40s)* |
| Timing of the program sessions | *"Uhm maybe time... it needs good timing; you know we are doing it for the sake of others who come from afar. That maybe shifting the afternoon program to be in the morning. ... in the afternoon, after you have eaten [lunch], the body gets kind of drunk... when the body is drunk, its actions are lazy-like. While in the morning, what does the body have? It has power. Yes, you can maybe do more exercises than that [than in the afternoon]." (P2F20s)* |
| Exercise design (intensity, diversity/variety, number, and duration of the exercise stations) | *"And also changing the duration [of a PR session] – I mean, you could have increased it, maybe to 1 and a half hour or even 2 hours, so that people's [participants'] bodies should remain relaxed [referring to the body relaxation benefit of PR] because most of them [participants], not to lie, maybe... they were doing these physical exercises only here – maybe they don't do it at home... I think you could have also added more [exercise] stations and other activities [different exercises] so that maybe people's bodies will relax..." (P9M20s)* |
| Self-management education | *".... you should have also been telling us... helping us... activities, "you can do this, do this, do this. When you arrive home, when you see [feel] this [symptoms/exacerbations], also do this." Because sometimes when I do it [exercise] at home, sometimes was like I was getting breathless with the exercise when I sit down. So, I don't know, what can we do in that case? Should we just use the medicine or whatever?" (P10F50s)* |

### Theme 5: Program continuation/maintenance at home

Motivated by PR benefits, participants were willing to continue exercising and/or being physically active at their homes, and some reported already doing so and even teaching others with similar lung conditions. However, most participants mentioned lack of equipment at home as a potential barrier to home-based exercises. On the other hand, potential enablers to home-based exercises and/or physical activities included using locally available equipment for the exercises or doing exercises that don’t require any equipment like walking or taking advantage of any available opportunities to stay physically active.

**Table 7:** Illustrative quotes of perspectives on home-based PR program.

| Sub-theme | Illustrative Quote |
| --- | --- |
| Willingness to continue program at home | *"Yes, I am ready to do everything in readiness to do the physical exercises. Or jogging, or lifting... I can lift like a packet of sugar for the physical exercises, I do. Jogging up a slight hill and down, I also do. Like the way I was doing here, I am doing likewise at home." (P1F40s)* |
| Motivators for continuing program at home | *"Yes, I would like [to continue the program at home] because it's helping me... I can do laundry now... yes, it [the program] is helping me. The body is not [no longer] weak [due to the program]." (P2F20s)* |
| Potential barriers to home-based program | *"Yes, I'm happy to continue the program at home but with fewer [exercise] stations, considering the equipment you use [in the hospital-based PR]; because we can't afford a treadmill. I may not be able to afford those weights [dumbbells]... So, continuing [exercising at home] can be there... but the resources to do so are the ones that may be missing. But I wish to continue, I can do it, but not according to the way you [PR delivery team] were doing it [at the hospital]." (P9M20s)* |
| Potential enablers to home-based program | *"But the other ones [some PR equipment], we can make, like those dumbbells... pouring sand, they told us, in bottles, it's possible. Sit to standing... that's also possible – some things were possible." (P10F50s)* |

## Discussion

Participants in this study reported negative experiences of living with CRD before participating in a PR program. Those negative experiences included poor physical health (including limitations in performing ADLs), poor psychological health (including anxiety), poor social health (including social exclusion and discrimination), poor sleep health (including sleep disturbance), and frequent inhaler use, hospital visits and admissions. Similar experiences were reported by people with CRDs in Sudan and Tanzania (17). After participating in the PR program, participants reported improved experiences of CRD in all the areas above, like those in Uganda (18).

CRDs pose an economic burden on patient and their households in Malawi. For example, chronic cough was found to be associated with unaffordable health-seeking costs among 608 people with CRDs (including post-TB lung disease, asthma, COPD, and bronchiectasis) in a cross-sectional community-based survey conducted in rural Malawi (19). These costs were mainly influenced by costs of transport and drugs. Therefore, the improvements in our participants’ physical activity performance and return to work, plus their reduced frequency of inhaler or medication use and hospital visits, may suggest a potentially positive economic impact of PR in Malawi for both patients and healthcare system. However, economic evaluation of PR in Malawi is yet to be conducted. In HICs, PR has proven to reduce patient and health system costs owing to reductions in exacerbations, medical visits, hospitalisation rates, and length of stay (20, 21).

The participants’ positive experiences of the PR program and some participants’ suggestion to include self-management education in the program to improve it suggest that PR improvements in CRD symptoms (e.g., breathlessness, cough, and body weakness), physical issues (e.g., a sense of physical wellbeing such as body relaxation and strength, performance of activities of daily living, and return to work), social issues (e.g., ability to interact with others and participate in social or group events), psychological issues (a sense of peace of mind, and reduction in anxiety), sleep quality, health literacy, medicine use, hospital visits and hospitalisation, might be important patient-reported outcomes for PR in the Malawian population but need further investigation such as through a core outcome set (COS) study.

Noteworthy, the PR delivery team in this study focused on delivering exercise training to participants during the PR sessions. Some participants’ suggestion in the current study that self-management education would have been included in the completed PR program echoes that suggests that health literacy or knowledge about their disease (including self-management) might be an important PR outcome in patients with CRDs in this setting. Self-management education is particularly recommended in the World Health Organisation’s package of essential rehabilitation interventions for patients with COPD (22). Our primary focus on delivering the exercise training component was based on evidence (largely from HICs) which presents exercise training as a cornerstone of PR that must prioritized (22) and, more recently, as one of the “essential” components of PR as it is underpinned by strong evidence (23). On the other hand, structured education, self-management training and smoking cessation support were found to be “desirable” (not essential) components of PR because strong evidence of their individual impacts is not yet available (23). Empirical investigation of the ideal PR composition in African settings such as Malawi is needed.

Barriers to PR commonly reported in LMICs and HICs include travel and transport difficulties (24, 25). Therefore, transport money provision was unsurprisingly mentioned by participants of the current study as one of the PR program’s success factors, facilitating their attendance and completion of the program. However, providing transport money to patients in Malawi is currently not a sustainable strategy to ensure long-term PR uptake at a hospital due to high economic challenges in the country (the current study was funded to provide participants with transport money, which is not the case in the routine clinical practice). Therefore, our finding reinforces the growing call for alternative models of PR delivery to be investigated, including home- and community-based PR, to address the travel and transport barrier, especially in Africa (24).

Encouragingly, participants of the current study expressed enthusiasm for home-based PR, and some were already continuing to exercise at home. Participants suggested that a safe and effective home-based PR program would be possible with a therapist-prescribed home exercise program, education on self-management skills, and use of locally available exercise equipment. In a randomised, controlled equivalence trial in Australia, a home-based PR program, using minimal resources (including readily available equipment in the patients’ home environment such as sit to stand from a dining chair and water bottles or bags of rice for upper limb weights) and little direct supervision, delivered short-term improvements in clinical outcomes (functional exercise capacity and health-related quality of life) that were equal to or greater than those seen in a conventional centre-based PR program in people with COPD (26). However, the effectiveness and participants’ experiences of a home-based PR model in LMICs, including Malawi, need to be empirically investigated as they may differ from HICs’.

Participants also mentioned good personal attributes of the PR delivery team (such as welcoming, heartwarming/warmful, cheerful, kind, polite, hardworking, and engaging or interactive) as one of the PR program’s success factors. These therapist attributes are likely to contribute to a strong therapeutic alliance, which is essential for increased patient compliance to treatment and subsequent improved patient outcomes (27, 28). However, the therapeutic alliance concept is yet to be extensively explored in the field of PR; few available PR studies have focused on patients’ (not therapists’) personality traits or characteristics (29, 30).

The inconvenient timing of PR sessions is a known barrier to attending PR (25). However, some participants’ suggestion to shift PR sessions from afternoon to morning hours for their convenience in this study highlights the current challenge of providing PR that meet individual patients’ needs in Malawi. The demand for rehabilitation services in the country is high. Yet, the rehabilitation centres (including at QECH, the study setting) are currently under-staffed, under-equipped, and less spacious to serve many patients efficiently. We chose afternoon hours for the PR sessions due to some of these reasons. We, therefore, join the participants’ call for further funding and government support towards rehabilitation services (including PR) if patients’ rehabilitation needs are to be met. This aligns with the World Health Organization “Rehabilitation 2030: a Call for Action” initiative (31).

### Study strengths and limitations

This qualitative study is part of the first feasibility and acceptability trial of PR in Malawi (32) and responds to other researchers’ calls for PR in Malawi (33, 34). But the theoretical transferability (35) of our findings is limited by the heterogeneity of study participants (for example, various CRDs, and comorbidities among participants including HIV infection) as well as the study having been conducted at a single site with a relatively small sample of patients. In addition, in the context of no provision of transport money, attendance and completion rates might differ. The translated participants’ quotes from Chichewa to English might have lost the original meaning (15). The interviews were conducted by TM and MMi who were also involved in the delivery of PR sessions; this might have influenced participants’ responses during the interviews. However, interview transcription and data analysis were led by the principal investigator (FMB) who had not been involved in both the delivery of the PR sessions and interviews.

## Conclusion

The PR program alleviated the participants’ perceived negative impacts of CRD on their physical, social, psychological and sleep health, associated with frequent inhaler use, hospital visits and admissions. In thinking about scaling PR within the Malawian context, key challenges need to be considered including PR financing, access to care in terms of transport and timing, task-shifting to primary care and community, and balancing equipment to optimize intervention quality with sustainability beyond the current PR program.

## Authors’ contributions

FMB, JR, and EC conceived and designed the study in 2018. FMB coordinated all activities of the study as the principal investigator, supervised by JR, EC, SBG, and FL. TM, BC, MMi, MMa, and BC screened and recruited patients with CRDs into the PR program. TM, BC, EC, MMi, CF, Mma, and LM delivered the PR sessions. TM and MMi interviewed the participants using a topic guide that benefited from the social science expertise of FL. FMB transcribed the interviews verbatim. FMB and TM analysed the data. FMB drafted the initial manuscript. MI, RNvZ-S, SH, MH, HS, EC, SBG, and FL critically reviewed the manuscript for important intellectual content and its overall preparation for publication. FMB revised the manuscript accordingly. All authors read and approved the final revised draft manuscript for journal submission and are all accountable for all aspects of the work.

## Data Availability

Raw data produced in the present study are available upon reasonable request to the authors.

